# Clinical characteristics and antibody response to SARS-CoV-2 spike 1 protein using the VITROS Anti-SARS-CoV-2 antibody tests in COVID-19 patients in Japan

**DOI:** 10.1101/2020.08.02.20166256

**Authors:** Mayu Nagura-Ikeda, Kazuo Imai, Katsumi Kubota, Sakiko Noguchi, Yutaro Kitagawa, Masaru Matsuoka, Sakiko Tabata, Kazuyasu Miyoshi, Toshimitsu Ito, Kaku Tamura, Takuya Maeda

## Abstract

**Background:** We evaluated clinical characteristics and the clinical utility of VITROS SARS-CoV-2 antibody tests according to COVID-19 severity in patients in Japan.

**Methods:** We analyzed 255 serum specimens from 130 COVID-19 patients and examined clinical records and laboratory data. Presence of total (IgA, IgM, and IgG) and specific IgG antibody for the spike 1 antigen of SARS-CoV2 was determined using VITROS Anti-SARS-CoV-2 antibody tests.

**Findings:** Overall, 98 (75.4%) and 32 (24.6%) patients had mild and severe COVID-19, respectively. On admission, 76 (58.5%) and 45 (34.6%) patients were positive for total and IgG antibody assays. Among 91 patients at discharge, 90 (98.9%) and 81 (89.0%) patients were positive for total and IgG antibody, respectively. Clinical background and laboratory findings on admission, but not the prevalence or concentration of total or IgG antibody, were associated with disease prognosis. Total and IgG antibody intensity were significantly higher in severe cases than in mild cases in serum collected after 11 days from onset, but not within 10 days.

**Conclusion:** VITROS Anti-SARS-CoV-2 Total and IgG assays will be useful as supporting diagnostic and surveillance tools and for evaluation of humoral immune response to COVID-19. Clinical background and laboratory findings are preferable predictors of disease prognosis.

## Introduction

The novel coronavirus disease 2019 (COVID-19), which is caused by severe acute respiratory syndrome coronavirus 2 (SARS-CoV-2) infection, was initially reported in December 2019 in Wuhan, China [1], and it has since gained pandemic status worldwide [2]. Serological tests are important because they provide faster results for surveillance and they support diagnosis based on the gold standard method of quantitative reverse-transcription polymerase chain reaction (RT-qPCR) [3]. Additionally, several reports have suggested that the antibody response to SARS-CoV-2 could be associated with disease severity [4-11]. Therefore, investigating this response in COVID-19 patients is important in order to assess the clinical utility of serological assays as supporting diagnostic tools and to understand the pathogenicity of SARS-CoV-2.

The serological tests currently available to detect total, IgM, and IgG antibodies against SARS-CoV-2 include enzyme-linked immunosorbent assay (ELISA), immunochromatography assay, and chemiluminescence immunoassay (CLIA) [12-15]. In particular, CLIA is widely used for COVID-19 detection in clinical settings because it is a rapid, high-throughput, and semi-quantitative method for detecting antibodies that is easy to perform. Commercial serological assays use a variety of SARS-CoV-2 antigens, including internal nucleocapsid protein (NP) and external spike (S) proteins. The S protein is composed of two subunits, S1 and S2. S1 contains the receptor binding domain (RBD), which is responsible for binding to the angiotensin-converting enzyme 2 (ACE2) receptor on the host cells at initiation of infection [16]. RBD is considered the major epitope recognized by SARS-CoV-2 neutralization antibodies [11, 17, 18]. Therefore, CLIA kits using S1 antigen are expected to be used both for diagnostic support and for the assessment of humoral immunity against SARS-CoV-2. The VITROS Anti-SARS-CoV-2 Total Antibody Test and VITROS Anti-SARS-CoV-2 IgG Antibody Test (Ortho Clinical Diagnostics, Raritan, NJ) both use the S1 protein of SARS-COV-2 and have been approved by the US Food and Drug Administration as commercial CLIAs for SARS-CoV-2 [19]. However, the clinical utility of these two antibody assays for COVID-19 and differences in antibody response in association with disease severity have not been well evaluated [20].

Here, we describe the clinical characteristics of patients with COVID-19 in Japan and report the clinical performance of and antibody response to the VITROS Anti-SARS-CoV-2 antibody assays for total (IgA, IgM, and IgG) and specific IgG antibodies to S1 protein of SARS-CoV-2, assessed in 255 serum samples collected from 130 COVID-19 patients with differing disease severity.

## Methods

### Patients with COVID-19 and serum samples

This study included 130 patients with COVID-19 who were referred to the Self-Defense Forces Central Hospital, a designated medical institution for specific infectious diseases in Tokyo, Japan, from February 26 to May 8, 2020. All were confirmed to have COVID-19 infection by RT-qPCR for SARS-CoV-2 according to the nationally recommended protocol [21]. We retrospectively collected patient information from electronic medical records. Serum samples were collected on the day of admission and during hospitalization. All serum samples were stored at −80°C before use in the antibody tests. This study was reviewed and approved by the Self-Defense Forces Central Hospital (approval number 01-011), and Saitama Medical University (approval number 917). Written informed consent was obtained from enrolled patient.

### Detection of antibodies for SARS-CoV-2

Total and IgG antibody assays against SARS-CoV-2 S proteins were performed using the VITROS Anti-SARS-CoV-2 Total Antibody Test and VITROS Anti-SARS-CoV-2 IgG Antibody Test (Ortho Clinical Diagnostics) according to the manufacturer’s instructions. Results were interpreted in accordance with these instructions as positive (signal for test sample /signal at cutoff [S/CO] ≥ 1.00) or negative (S/CO < 1.0) for both total and IgG assays.

### Definition of cases

Symptomatic cases were then subdivided into severe and mild groups. Severe symptomatic cases were defined as patients with clinical symptoms of pneumonia (saturation of percutaneous oxygen [SpO2] <93% and need for oxygen therapy). Other symptomatic cases were classified as mild. The criterion for discharging COVID-19 patients from hospital was a negative result on two consecutive RT-qPCR tests [21] using throat swab specimens under the Infectious Disease Control Law in effect from February 11 to May 13, 2020.

### Statistical analysis

Continuous variables are expressed as the mean ± standard deviation (SD) or median interquartile range [IQR] and were compared using the t-test or Wilcoxon Rank Sum test for parametric or non-parametric data, respectively. Categorical variables are expressed as number (%) and were compared using the χ2 test or Fisher’s exact test. All statistical analyses were performed using R (v 4.0.0; R Foundation for Statistical Computing, Vienna, Austria [http://www.R-project.org/]).

## Results

### Clinical characteristics of patients with COVID-19 in Japan

Table 1 shows the baseline clinical characteristics of all 130 patients. Briefly, the patients ranged in age from 18 to 91 years (median, 45 years; IQR, 37–59 years); 77 (59.2%) were male and 53 (40.8%) were female. The time from symptom onset to admission day was 3–21 days (median, 8 days; IQR, 6–10 days). Among the 66 (50.8%) patients who had underlying diseases, hypertension (17.7%) and respiratory disorder (17.7%) were most frequently observed. The most common clinical features on admission were fever (91.5%) and cough (53.1%). Furthermore, 76 (58.5%) and 45 (34.6%) patients were positive for total and IgG antibody assay, respectively, on admission. In total, 126 (96.9%) patients recovered and were discharged from hospital, and 4 (3.1%) patients died during the observation period. The median duration from initial symptom onset to confirmatory twice-negative PCR for SARS-CoV-2 RNA was 17 days (IQR 14-22 days). Also, 32 (24.6%) patients required supplemental oxygen therapy and were classified as having severe COVID-19. The medium period from symptom onset to requiring supplemental oxygen therapy was 9 days (IQR 7-11 days). Among the 32 severe cases, high-flow nasal cannula therapy was required in 14 (10.8%) and mechanical ventilation was required in 5 (3.8%).

**Table 1.** Clinical characteristics of patients with COVID-19.

|  | **All patients**<br>**(N=130)** | **Mild**<br>**(n=98)** | **Severe**<br>**(n=32)** | ***p*-value** |
| --- | --- | --- | --- | --- |
| **Background characteristics** |  |  |  |  |
| Age (years) | 45<br>[37-59] | 44<br>[35-53] | 62<br>[57-69] | < 0.001 ** |
| Time from onset to admission (days) | 8<br>[6-10] | 8<br>[6-10] | 8<br>[6-9] | 0.327 |
| Sex |  |  |  | 0.002 * |
| Female | 53 (40.8) | 47 (48.0) | 6 (18.8) | - |
| Male | 77 (59.2) | 51 (52.0) | 26 (81.3) | - |
| Smoking history |  |  |  | 0.411 |
| Yes | 61 (46.9) | 43 (43.9) | 18 (56.3) | - |
| No | 69 (53.1) | 55 (56.1) | 14 (43.8) | - |
| Underlying diseases |  |  |  |  |
| Total | 66 (50.8) | 39 (39.8) | 27 (84.4) | < 0.001 ** |
| Cardiovascular disease | 9 (6.9) | 6 (6.1) | 3 (9.4) | 0.688 |
| Hypertension | 23 (17.7) | 10 (10.2) | 13 (40.6) | < 0.001 ** |
| Diabetes | 5 (3.8) | 1 (1.0) | 4 (12.5) | 0.013 * |
| Respiratory disorder | 23 (17.7) | 14 (14.3) | 9 (28.1) | 0.107 |
| Malignancy | 3 (2.3) | - | 3 (9.4) | 0.014 * |
| **Clinical features on admission** |  |  |  |  |
| Fever | 119 (91.5) | 88 (89.8) | 31 (96.9) | 0.412 |
| Cough | 69 (53.1) | 50 (51.0) | 19 (59.4) | 0.541 |
| Malaise | 39 (30.0) | 32 (32.7) | 7 (21.9) | 0.371 |
| Diarrhea | 31 (23.8) | 26 (26.5) | 5 (15.6) | 0.335 |
| Headache | 39 (30.0) | 33 (33.7) | 6 (18.8) | 0.179 |
| Dyspnea | 15 (11.5) | - | 15 (46.9) | - |
| Tachypnea | 15 (11.5) | - | 15 (46.9) | - |
| **Laboratory findings on admission** |  |  |  |  |
| AST (IU/L) | 30<br>[22-43] | 27<br>[20-33] | 55<br>[36-78] | < 0.001 ** |
| ALT (IU/L) | 31<br>[18-45] | 27<br>[16-41] | 39<br>[34-66] | < 0.001 ** |
| LDH (IU/L) | 231<br>[182-323] | 208<br>[168-267] | 376<br>[304-506] | < 0.001 ** |
| Sodium concentration | 139<br>[136-141] | 140<br>[138-142] | 136<br>[134-138] | < 0.001 ** |
| C-reactive protein (mg/dL) | 1.3<br>[0.2-4.6] | 0.6<br>[0.1-2.3] | 9.0<br>[4.6-13.9] | < 0.001 ** |
| White blood cell count (/μL) | 5,152<br>[3,808-6,440] | 4,878<br>[3,692-6,112] | 6,015<br>[3,354-8,355] | 0.007* |
| Platelet count (×10<sup>4</sup>/μL) | 19.5<br>[15.7-26.6] | 19.8<br>[15.9-26.7] | 22.8<br>[15.5-23.8] | 0.658 |
| Lymphocyte count | 1,054<br>[774-1447] | 1,206<br>[896-1,692] | 698<br>[501-875] | < 0.001** |
| Neutrophil/lymphocyte ratio | 2.8<br>[1.8-4.7] | 2.4<br>[1.4-3.5] | 7.3<br>[3.8-11.6] | < 0.001** |
| **Antibody tests on admission** |  |  |  |  |
| Total antibody, positive | 76 (58.5) | 60 (61.2) | 16 (50.0) | 0.304 |
| Total antibody (S/CO) | 3.71<br>[0.2-33.1] | 4.25<br>[0.1-32.9] | 1.90<br>[0.4-45.6] | 0.152 |
| IgG antibody, positive | 45 (34.6) | 34 (34.7) | 11 (34.4) | 1.000 |
| IgG antibody (S/CO) | 0.1<br>[0-3.1] | 0<br>[0-3.6] | 0.1<br>[0-2.5] | 0.322 |
| **Treatment** |  |  |  |  |
| Antibiotics | 16 (12.3) | - | 16 (50.0) | - |
| Lopinavir/ritonavir | 2 (1.5) | - | 2 (6.3) | - |
| Favipiravir | 11 (8.5) | 2 (2.0) | 9 (28.1) | - |
| Methylprednisolone | 13 (10.0) | - | 12 (37.5) | - |
| Anticoagulant drugs | 12 (9.2) | - | 12 (37.5) | - |
| Oxygen therapy | 32 (23.6) | - | 32 (100) | - |
| HFNC | 14 (10.8) | - | 14 (43.8) | - |
| NPPV | 1 (0.8) | - | 1 (3.1) | - |
| IMV | 5 (3.8) | - | 5 (15.6) | - |
| **Outcome** |  |  |  |  |
| Recovery | 126 (96.9) | 98 (100) | 28 (87.5) | - |
| Death | 4 (3.1) | - | 4 (12.5) | - |

Data are presented as n (%), or median [interquartile range], unless otherwise specified.
\* $p < 0.05$ , \*\* $p < 0.001$ .
AST, aspartate aminotransferase; ALT, alanine aminotransferase; LDH, lactate
dehydrogenase; S/CO, sample/signal at cutoff; HFNC, high-flow nasal cannula; NPPV,
noninvasive positive pressure ventilation; IMV, intermittent mandatory ventilation.

Univariate analysis revealed that age (*p* < 0.001) and male preponderance (*p* = 0.002) were significantly higher in the severe COVID-19 group than in the mild COVID-19 group. The prevalence of underlying disease was also higher in severe cases (*p* < 0.001), with significant differences especially in the prevalence of hypertension (*p* < 0.001), diabetes mellitus (*p* = 0.013), and malignancy (*p* = 0.014). There were no significant differences in clinical features between the two groups. Blood test results revealed severe cases had higher concentrations of aspartate aminotransferase (AST), alanine aminotransferase (ALT), lactate dehydrogenase (LDH), and C-reactive protein (CRP), higher white blood cell count and neutrophil/lymphocyte ratio (NLR), and lower sodium level and lymphocyte count than mild cases. There were no significant differences in the positivity rate or S/CO for either total or IgG antibody assay on admission between the two groups.

All 91 patients had paired serum samples, collected on admission and discharge (Table 2), and among them on discharge, 90 (98.9%) and 81 (89.0%) patients were positive for total and IgG antibody, respectively. The S/CO of total and IgG antibody on discharge was significantly higher in severe cases than in mild cases (*p* < 0.001 for total and *p* = 0.009 for IgG antibodies), while there was no significant difference in the seropositive rate for either total or IgG antibody between the two groups on discharge (*p* > 0.05).

**Table 2.** Rate of positive total and IgG antibody assays on admission and discharge.

|  |  | Positivity rate |  |  | S/CO |  |  |
| --- | --- | --- | --- | --- | --- | --- | --- |
|  |  | On admission | On discharge | *p*-value | On admission | On discharge | *p*-value |
| **Total Ab** | **N = 91** | 54<br>(59.3%) | 90<br>(98.9%) | < 0.001** | 3.7<br>[0.1 - 33.1] | 66.7<br>[32.3 - 124.3] | < 0.001** |
|  | **Mild = 65** | 39<br>(60.0%) | 64<br>(98.5%) | < 0.001** | 4.3<br>[9.1-32.9] | 45.9<br>[19.1-108] | < 0.001** |
|  | **Severe = 26** | 15<br>(57.7%) | 26<br>(100.0%) | < 0.001** | 1.9<br>[0.4 - 30.0] | 106.6<br>[60.8 - 172.5] | < 0.001** |
|  | ***p*-value** | 1.000 | 1.000 |  | 0.805 | < 0.001** |  |
|  |  | Positive rate |  |  | S/CO |  |  |
|  |  | On admission | On discharge | *p*-value | On admission | On discharge | *p*-value |
| **IgG Ab** | **Total = 91** | 34<br>(37.4%) | 81<br>(89.0%) | < 0.001** | 0.1<br>[0 - 3.1] | 8.3<br>[3.4 - 10.8] | < 0.001** |
|  | **Mild = 65** | 24<br>(36.9%) | 56<br>(86.2%) | < 0.001** | 0.1<br>[0 - 3.6] | 6.2<br>[2.1 - 5.9] | < 0.001** |
|  | **Severe = 26** | 10<br>(38.5%) | 25<br>(96.2%) | < 0.001** | 0.4<br>[0 - 2.3] | 10.8<br>[8.7 - 12.6] | < 0.001** |
|  | ***p*-value** | 1.000 | 1.000 |  | 0.992 | 0.009 * |  |

Data are presented as n (%) or median [interquartile range], unless otherwise specified.
Ab, antibody; S/CO, sample/signal at cutoff
\* $p < 0.05$ , \*\* $p < 0.001$ .

### Comparison of antibody response in COVID-19 patients

For the 255 collected serum specimens obtained from 130 patients, the period from initial symptom onset to sample collection was 1–49 (median 13 days, IQR 9-18). Among the serum specimens, the positivity rate for total and IgG increased with longer clinical course, and total antibody was detected earlier than specific IgG antibody (Figure 1). In mild and severe cases, total antibody assay was positive in 54.4% (37/68) and 47.8% (11/23) of specimens collected within 10 days of onset, in 95.6% (86/90) and 100% (28/28) collected within 11–20 days, and 100% (23/23) and 100% (23/23) collected >21 days after onset, respectively. The corresponding detection rates for IgG antibody were 25.0% (17/68) and 26.1% (6/23), 75.6% (68/90) and 89.3% (25/28), and 100% (23/23) and 100% (23/23). There was no significant difference in the prevalence of total or IgG antibody positivity between severe and mild cases (*p* > 0.05). Among the 130 patients, 3 serum samples collected from 2 patients were negative for total antibody but positive for IgG antibody. Among both mild and severe cases, S/CO also increased with a longer clinical course (Figure 2). S/CO for the total and IgG antibody assays was significantly higher in severe cases than in mild cases among serum samples collected from patients 11–20 days after onset (*p* < 0.001 for both total and IgG antibodies) and >21 days after onset (*p* =0.039 for total and *p* < 0.001 for IgG antibodies), but there was no significant difference within 10 days (*p* > 0.05 for both total and IgG antibodies; Figure 3).

**Figure 1.**
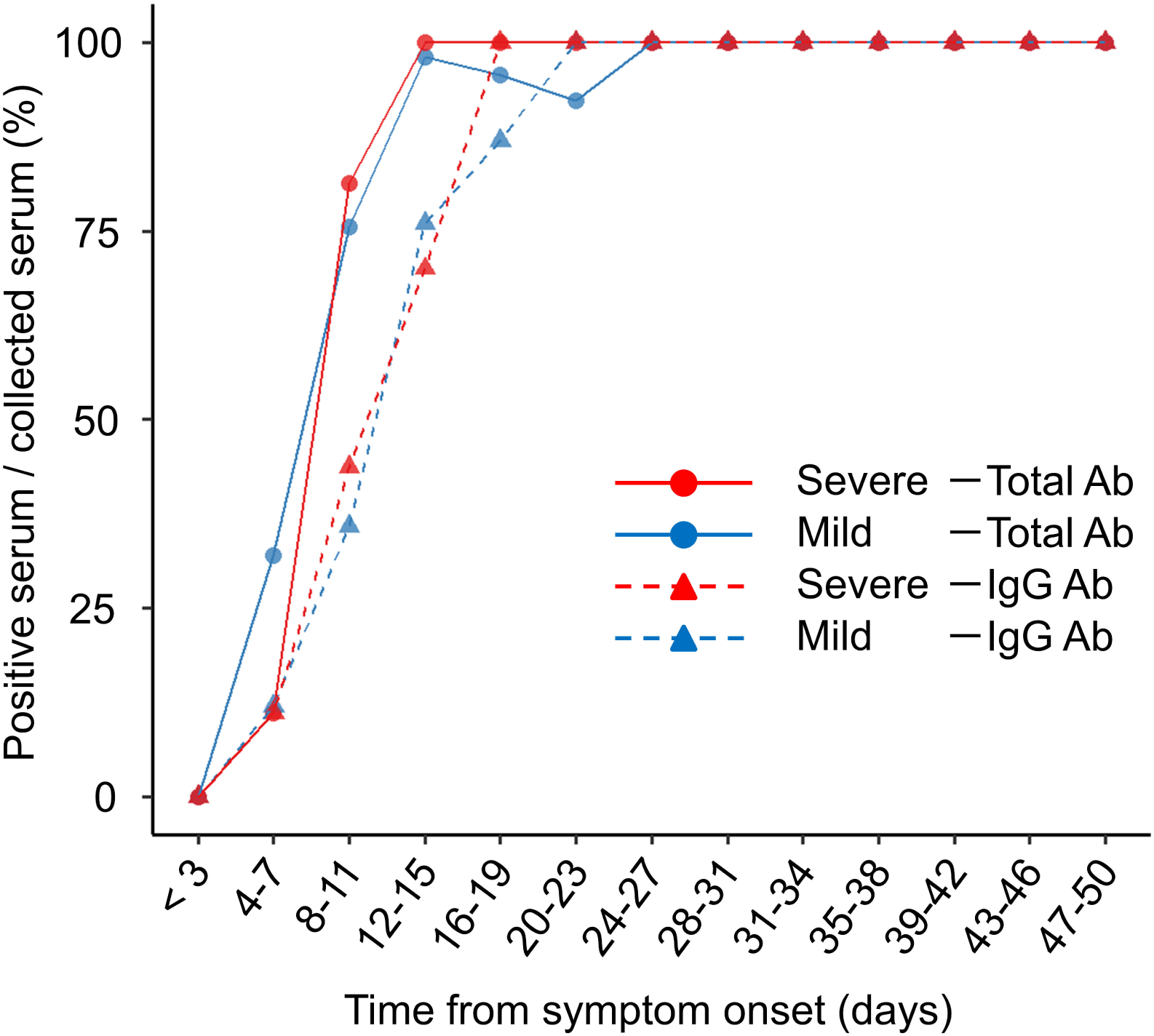
Seropositive rate of total and IgG antibody response in serum specimens collected from mild and severe COVID-19 cases after symptom onset. Plot shows seropositive rate for total and IgG antibody assays per total serum collected from symptom onset. Solid and dashed lines indicate total and IgG antibody assays, respectively. Blue and red lines indicate mild and severe cases, respectively. Ab, antibody.

**Figure 2.**
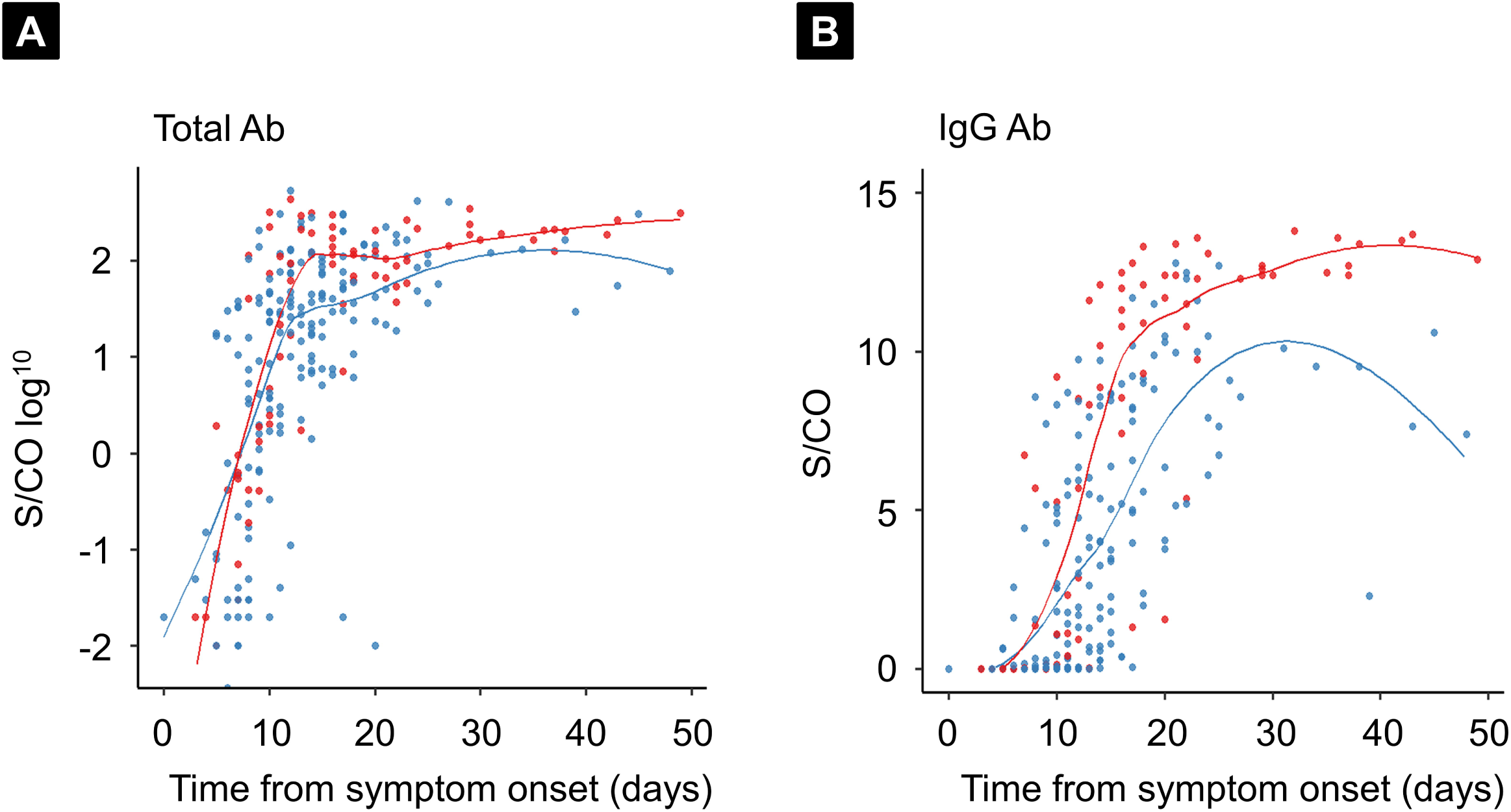
Kinetics of total and IgG antibody response between mild and severe cases. Plots show time to sample collection from symptom onset and signal for test sample/signal at cutoff [S/CO] for total and IgG antibodies. (A) Total antibody, (B) IgG antibody. Blue and red lines indicate regression curves of mild and severe cases, respectively. Ab, antibody.

**Figure 3.**
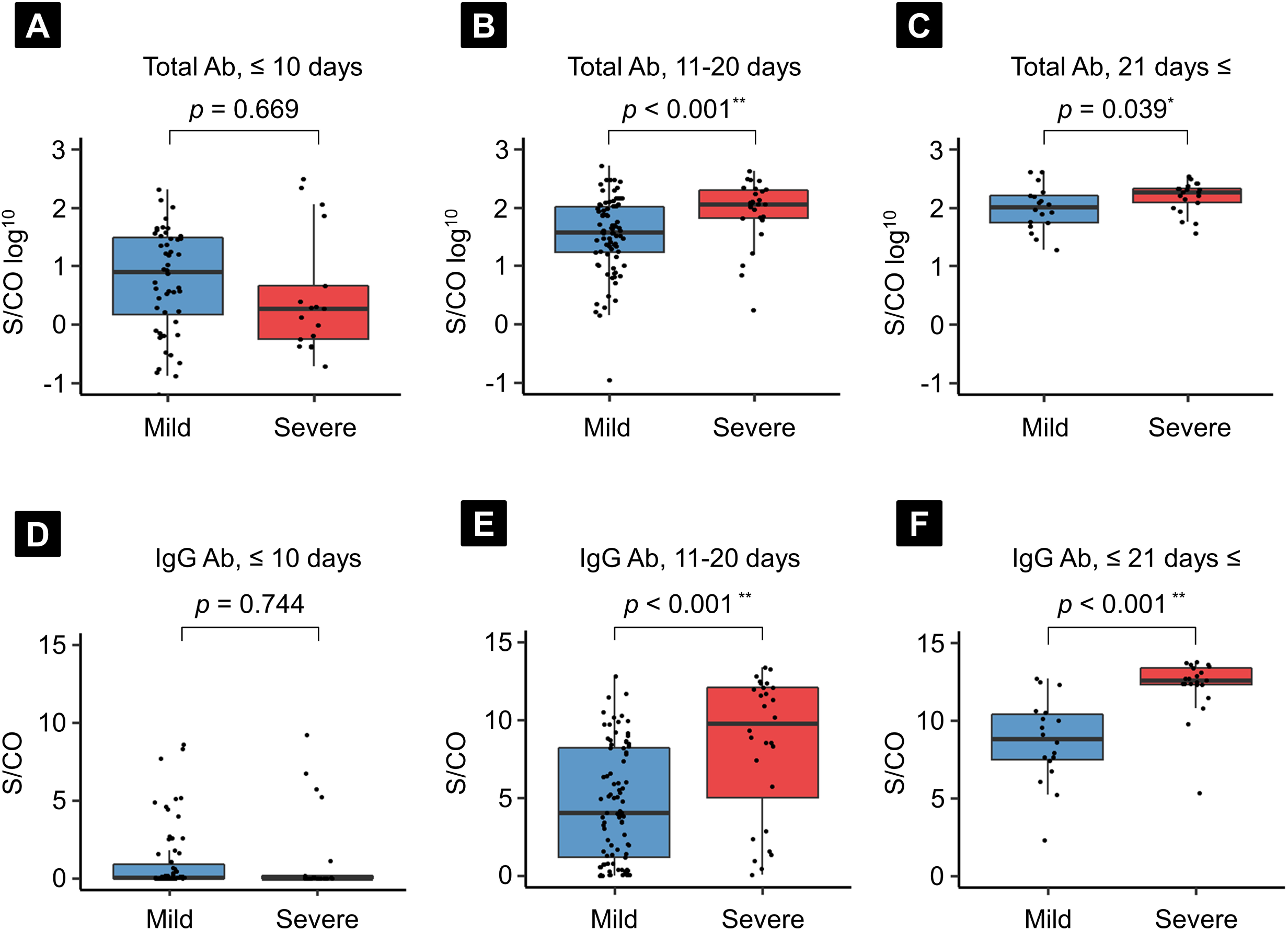
Comparison between mild and severe cases of total and IgG antibody response for total antibody (A-C) and IgG antibody (D-E) in serum specimens collected at different timings from symptom onset. Correlation coefficient calculated using Wilcoxon’s rank-sum test. * *p* < 0.05, ** *p* < 0.001. Ab, antibody.

## Discussion

Here, we have presented the clinical characteristics of patients with COVID-19 in Japan and provided evidence for the clinical utility of the VITROS Anti-SARS-CoV-2 Total and IgG assays in diagnosing COVID-19 as well as for differences in antibody response against the SARS-CoV-2 S1 protein with regard to disease severity.

Total antibody (IgA, IgM, and IgG) for SARS-CoV-2 S1 protein was detected earlier than IgG antibody, and therefore total antibody assays appear to be preferable for initial diagnostic testing for COVID-19 in patients who develop symptoms within 2 weeks. The sensitivity of the total antibody assay was 47.8-54.4% and 95.6-100% in serum collected within 10 days and >11 days from onset, respectively. This finding was consistent with previous reports, which showed rapidly increased detection of IgA, IgM, and IgG antibodies against S protein from day 10-15 after onset based on ELISA [22, 23], and other commercial CLIA kits that used the NP or S1+2 protein as an antigen [24-26]. In this study, we demonstrated that disease severity did not affect the positivity rate of the VITROS Anti-SARS-CoV-2 Total antibody assay. Therefore, the total antibody assay will be useful in clinical settings as a supporting diagnostic tool alongside the gold standard RT-qPCR for both mild and severe cases.

Regarding the timing of discharge with negative viral RNA results from throat swab specimens, the positivity rate for total and IgG antibodies was 98.9% and 89.0%, respectively. Additionally, almost 100% of serum samples were positive for total and IgG antibodies in the late phase of the clinical course (>21 days) in both mild and severe cases. Generally, both IgA and IgM antibodies have a relatively shorter lifespan than IgG antibody. Also, IgA antibody for S1 protein is reported to be less specific than IgG [11]. Taken together, the IgG antibody assay is preferable to the total antibody assay for surveillance of COVID-19 seroprevalence. Liu et al. reported a strong correlation of the IgG titer against S1 with the RBD. [27] Although further study is still needed to clarify the correlation of antibody titer determined by the VITROS Anti-SARS-CoV-2 antibody assays and neutralization antibody, the weight of evidence to date suggests that the VITROS Anti-SARS-CoV-2 antibody assays also may be used for the evaluation of humoral immunity against SARS-CoV-2.

In terms of the discrepancy in antibody response to S protein with regard to severity, Zhao et al. reported a significantly higher antibody titer for S protein in severe cases than mild cases based on ELISA [22]. Our findings of higher S/CO for both the total and IgG antibody assays in severe cases in serum collected after 11 days from symptom onset are consistent with theirs. However, antibody titer within 10 days after onset or on the day of admission did not differ significantly according to disease severity. In our study, median time from symptom onset to admission and time from onset to need for oxygen therapy was 8 and 9 days, respectively. Thus, antibody titer may be an indicator of disease severity, but it may not be a predictor of prognosis in the early phase. In this study, clinical background (distribution of age, sex, and underlying disease) and laboratory findings (AST, ALT, LDH, CRP, sodium, WBC count, lymphocyte count, and NLR) on admission were associated with disease prognosis, as reported previously [28-32]. Therefore, it is preferable to use these clinical background characteristics and laboratory findings as predictors of disease prognosis in the early phase of the clinical course.

This study has some limitations. First, despite the confirmed high specificity of VITROS Anti-SARS-CoV-2 antibody assays [19, 20], specificity should be further analyzed using clinical specimens from non-COVID-19 patients. Also, recent reports showed a time-dependent decrease in antibody titer after the initial infection, especially in asymptomatic and mild cases [33]. Further studies are warranted to determine the utility of VITROS Anti-SARS-CoV-2 antibody assays as diagnostic and surveillance tools for COVID-19.

## Conclusion

The qualitative results of the VITROS Anti-SARS-CoV-2 Total and IgG assays are useful, as are their S/CO values in supporting diagnosis and surveillance in both mild and severe COVID-19 cases. There are differences in antibody response to SARS-CoV-2 S1 protein according to disease severity, and thus clinical background characteristics and laboratory findings are preferable predictors of disease prognosis in the early phase of the clinical course.

## Data Availability

The raw data used in this study is available from the corresponding author upon reasonable request.

## Conflict of interest

The authors declare that they have no conflicts of interest.

## Funding

This research did not receive any specific grant from funding agencies in the public, commercial, or nonprofit sectors.

## Acknowledgments

We thank the clinical laboratory technicians at the Self-Defense Forces Central Hospital for sample collection, and all members of the COVID-19 Task Force at the Self-Defense Forces Central Hospital and participating members drawn from other institutes of the Japan Self-Defense Forces.

## Authors’ contributions

TM and KI: study conception and design; MNI, KI, and ST: clinical data collection and data analysis; SN, YK, and MM: experiment data analysis; MNI, KI, and TM manuscript drafting and editing; SN, YK, MM, TI, and KK: manuscript revision; TI and KK study supervision. All authors have read and approved the final manuscript.

## Notes

### Competing Interest Statement

The authors have declared no competing interest.

### Author Declarations

This study was reviewed and approved by the Self-Defense Forces Central Hospital (approval number 01-011), and Saitama Medical University (approval number 917). Written informed consent was obtained from enrolled patient.

